# Sex-based differences in imaging-derived body composition and their association with clinical malnutrition in abdominal surgery patients

**DOI:** 10.1101/2025.04.05.25325276

**Authors:** Raheema Damani, Shubha Vasisht, Valerie Luks, Gracia Vargas, Charlene Compher, Paul M. Titchenell, Gregory Tasian, Hongzhe Li, Gary D. Wu, Walter R. Witschey, Victoria M. Gershuni

## Abstract

**Structured Abstract:** *Background:* Malnutrition significantly impacts surgical outcomes yet is difficult to identify preoperatively. Few studies have investigated the association between comprehensive body composition assessment and malnutrition in males and females separately. This study evaluates sex-specific associations between preoperative imaging-derived body composition features and malnutrition in abdominal surgery patients.

*Methods:* We retrospectively analyzed patients who underwent computed tomography (CT) scans and elective abdominal surgery at a single institution (2018-2021). Preoperative CT scans were assessed using deep learning to quantify five muscle groups and two fat depots. Malnutrition was diagnosed by registered dietitians using standardized criteria. Sex-specific associations with malnutrition were evaluated using logistic regression.

*Results:* Among 1,143 patients (52% female), clinical malnutrition was diagnosed in 20.2% of patients, with prevalence varying by procedure type (3.5-38.2%). Malnutrition was associated with reduced muscle volume for both sexes; in contrast, malnutrition was associated with myosteatosis in 3 of 5 muscle groups for females only. In males, malnutrition was associated with decreased psoas volume (OR 0.59 SD, p<0.01), decreased quadratus lumborum volume (OR 0.59 SD, p<0.01), and reduced erector spinae attenuation (OR 0.66 SD, p=0.048). In females, decreased psoas volume (OR 0.55 SD, p<0.001) and attenuation (OR 0.64 SD, p<0.01) were associated with malnutrition. Both sexes demonstrated increased subcutaneous fat attenuation associated with malnutrition (males: OR 1.51 SD, p<0.01; females: OR 1.73 SD, p<0.001), while increased visceral fat attenuation (OR 1.4 SD, p=0.027) was associated with malnutrition only in females.

*Conclusions:* Males and females differ in baseline body composition and features associated with clinical malnutrition. Comprehensive deep learning analysis of muscle and fat characteristics from cross-sectional imaging provides insight into the sex-specific relationships between body composition and malnutrition in the preoperative setting and provides an opportunity for early identification of patients with greater nutrition-related surgical risk.

## INTRODUCTION

Malnutrition is an independent risk factor for poor outcomes after abdominal surgery. Surgical patients with malnutrition have increased perioperative morbidity, mortality, hospital readmission, and length of stay. (1) Malnutrition is highly prevalent yet underrecognized, partly due to lack of objective and easy to implement methods for nutritional assessment. While malnutrition is a modifiable risk factor, it is hard to identify using current methods, which heavily rely on clinical assessment by a trained registered dietitian. Efforts to improve preoperative diagnosis of malnutrition have thus far been stymied by the lack of a reliable, automated assessment tool that can be implemented in a high-throughput manner.

Muscle mass has been highlighted as an important component of nutrition assessment and a phenotypic indicator of nutritional status according to the Global Leadership Initiative on Malnutrition criteria. Muscle mass corresponds with physiologic reserve and the ability to respond to stress, including surgery. Independent of fat mass and body mass index, loss of muscle mass (sarcopenia), has been shown to be a predictor of metabolic disease, mortality, (2) and adverse outcomes after surgery. (3-10) More recently, fatty replacement of muscle tissue (myosteatosis), which includes both intermuscular fat accumulation and intramyocellular lipid deposition, has been linked to impaired muscle function, disuse atrophy, aging and poor nutritional status. (11-14) Variation in intramyocellular lipid deposition has been linked to diet and exercise with increased intramyocellular lipids in response to prolonged starvation. (15, 16) Myosteatosis is seen with insulin resistance, cardiometabolic disease, and metabolic dysfunction, independent of obesity or excess weight. (17)

Body composition analysis using clinically obtained computed tomography (CT) scans provide detailed measurements of both skeletal muscle and fat stores, offering an objective method to assess nutritional status in a potentially more sensitive manner than traditional risk factors and screening tools. (18) Using CT, the size and quality of multiple tissue types can be quantified, including muscle size, fatty replacement of muscle tissue, visceral fat, and subcutaneous fat. Single-slice measures of cross-sectional area of muscle (skeletal muscle index, SMI) at the third lumbar vertebrae (L3) have been explored for potential to predict malnutrition in surgical populations. (10, 19, 20) However, males and females have known sex-specific differences in muscle size, deposition, and quality; thus, reliance on SMI alone may underestimate risk of malnutrition in females. (21-23) Females have smaller muscle size and decreased muscle quality (greater fatty replacement) compared to males due to hormonal regulation, and they lose muscle at a slower rate than males during periods of stress. (23, 24) These sex-specific characteristics may lead to an underestimation of malnutrition risk among females when assessed using muscle size alone.

Incorporation of more detailed body composition, including visceral and subcutaneous fat, allows for greater understanding of the complex relationship between these metabolically active and hormone-regulated depots and the pathophysiologic state of malnutrition. Recent evidence suggests that decreased visceral fat with increased attenuation on imaging is associated with worse oncologic outcomes and worse survival in head and neck cancer, which has notoriously high rates of malnutrition. (26) Sex-specific differences in body composition, specifically visceral fat accumulation seen predominantly in males with obesity are linked to worse cardiometabolic outcomes and inflammation. (23, 25) However, the sex-specific differences between the relationship of malnutrition and hormone-mediated fat depot utilization are not clear. There is a critical gap in our understanding of the association between body composition features and nutritional status in males and females.

This study aims to address this knowledge gap by evaluating the associations between imaging derived body composition features and malnutrition among males and females undergoing abdominal surgery. Here we apply a novel, volumetric approach using an automated machine learning algorithm to measure five distinct muscle groups and two fat depots on clinically obtained preoperative abdominal CT scans. (27, 28) The algorithm quantifies both the size and quality of each muscle and fat depot, offering a more comprehensive evaluation of body composition than L3 single-slice cross sectional measures. (19, 20, 27) We describe the imaging features of body composition for males and females in an abdominal surgery cohort and then determine the sex-specific association between imaging features of body composition and the likelihood of diagnosis of clinical malnutrition.

## METHODS

### Study Population

We conducted a retrospective cross-sectional study that included adult patients at a single quaternary care institution between 2018 to 2021 who underwent elective abdominal surgery and had a routine preoperative CT scan performed within 90 days before surgery. (29) Data was prospectively collected for standardized inclusion in the American College of Surgeons National Surgery Quality Improvement Program (NSQIP) registry for five distinct procedures (pancreatectomy, hepatectomy, colectomy, proctectomy, and ventral hernia repair). (30) Additional clinical, demographic, and comorbidity data, including serum albumin, weight, height, body mass index (BMI), and clinical nutrition assessment variables were abstracted from the electronic health record (Epic Systems).

### Nutrition Outcomes

Nutritional status assessment was performed after admission to the hospital according to standard of care as outlined by the Academy ASPEN Indicators of Malnutrition (AAIM). (31) This nutrition care process includes two steps: (1) screening within 24 hours of admission to the hospital using the validated Malnutrition Screening Tool (32) and (2) clinical assessment by a Registered Dietitian required for the formal diagnosis of clinical malnutrition. The AAIM assessment includes degree of weight loss over time and loss of muscle and fat stores on physical exam. (31) Patients who do not screen "At Risk" in the first step do not proceed to the formal Registered Dietitian assessment and are assumed to have adequate nutritional reserve. Patients with no screening assessment were treated as having no risk for malnutrition. The primary outcome of this study was diagnosis of clinical malnutrition as determined by the Registered Dietitian evaluation, which by protocol occurs in the immediate post-operative period during hospital admission.

### Computed Tomography Acquisition and Deep Learning-based Image Analysis

Patients with a clinically obtained preoperative CT scan performed within 90 days prior to admission were included. Imaging was performed using multidetector CT scanners from a variety of vendors (Philips Healthcare, GE Healthcare, Siemens Healthineers, Canon Medical Systems, Toshiba). The tube current and iodinated contrast application (including phase of contrast) varied based on the clinical setting. The slice thickness varied between 2-5 mm. The tube voltage varied between 80 and 140 kVp (median: 100 kVp, IQR: 20 kVp). Scans that were not collected in axial orientation with isotropic in-plane resolution or contained an inferior-to-superior field-of-view of less than 100 mm, or slice thickness outside of 2-5 mm range, were excluded from analysis.

Quantification of L3 single-slice measures was performed automatically using licensed software from Data Analysis Facilitation Suite version 3.6 (Voronoi Health Analytics Inc, https://www.voronoihealthanalytics.com/dafs). Previously validated L3 single-slice cross-sectional area measures included skeletal muscle index (SMI, cm^2^/m^2^) and skeletal muscle radiation attenuation (SMRA, HU).(19, 33, 34) Volumetric quantification was performed using a novel deep learning approach previously described by our group. (27, 28) The abdominal cavity was delineated automatically as the region between the inferior aspect of the lung (T12 vertebral level) and the inferior aspect of the L5 vertebrae. Segmentations included automated quantification of volume and attenuation of five individual abdominal muscle groups (psoas, erector spinae, quadratus lumborum, lateral abdominals, and rectus abdominus) and two fat depots (visceral and subcutaneous). Muscle quality was determined by the attenuation of voxels contained within the muscle contour which included both muscle tissue and intermuscular adipose. Once the abdominal contour was segmented, fat voxels were identified using attenuation thresholds of -190 to -30 HU. Voxels within the abdominal contour were labeled as visceral fat, and voxels outside the abdominal contour were labeled as subcutaneous fat. All volume measurements were adjusted for patient height squared to generate a volume index (cm^3^/m^2^). Due to the novel nature of these volumetric measurements, sex-specific z-score normalization was performed using the sex-specific mean and standard deviation derived from this cohort. A list of all volumetric and single-slice imaging features can be found in Supplemental Table 1.

### Statistical Analysis

Descriptive statistics were used to summarize the surgical cohort stratified by sex. Continuous variables were reported as mean and standard deviation (SD) or median and interquartile range (IQR) based on distribution of data and analyzed using Student’s t-test or Mann-Whitney U test, respectively. Categorical variables were reported with absolute and relative frequencies and compared using Chi-square or Fisher’s exact test. Principal component analysis was used to visualize the spatial distribution of body composition features among males and females. (35) Permutational Multivariate Analysis of Variance (PERMANOVA) was used to determine if the multivariate means (group centroid) were significantly different between males compared to females. (36) The Permutation Test for Homogeneity of Multivariate Dispersion (PERMDISP) was performed to assess whether there was significant dispersion of values around the centroid for males compared to females. (37)

Logistic regression (38) models were built to estimate the relationship between imaging derived body composition features and clinical malnutrition, adjusting for age, weight, race, ASA classification and tobacco use. (22, 39) Odds ratios (OR) with 95% CI were computed for each feature for males and females separately.

All statistical analyses were performed in R (R Core Team, version 4.1.2; Foundation for Statistical Computing, Vienna, Austria). A p-value of less than 0.05 was considered statistically significant. This study was carried out in compliance with all relevant guidelines and regulations (TRIPOD-AI) and was determined to be exempt from informed consent by the institutional review board due to deidentified and retrospective nature of dataset.

## RESULTS

The final surgical cohort (n=1143) with available preoperative imaging was comprised of 48% males and 52% females. The mean age was 60.5 ± 14.6 years with an average body mass index (BMI) of 28.7 ± 7.1 kg/m^2^. 75% were white and 65.7% had an ASA score greater than 3. The most common procedure type was colectomy (504) followed by ventral hernia repair (229), pancreatectomy (204), hepatectomy (155), and proctectomy (51); 65.4% of surgeries were performed via an open approach. No clinically meaningful differences between sexes were observed for age, race, BMI, tobacco use, procedure type, operative approach, albumin, MST score, or incidence of clinical malnutrition. Males had significantly greater weight (86.1 vs. 73.6 kg, p<0.001), ASA class (p=0.007), prevalence of diabetes (21 vs. 14.5%, p=0.008) and hypertension (49.4 vs. 43.4%, p=0.04) compared to females. There were no significant differences between males and females for post-operative complications or prolonged length of stay (Table 1).

**Table 1.** Demographic and Clinical Characteristics for Abdominal Surgery Patients segregated by Sex.

| Characteristic | Total<br>(n=1143) | Male<br>(N=544) | Female<br>(N=599) | p-value |
| --- | --- | --- | --- | --- |
| Age (years) | 60.51 ± 14.64 | 60.9 ±14.8 | 60.9 ±15 | 0.931 |
| Race |  |  |  |  |
| White | 856 (74.9%) | 419 (77%) | 437 (73.3%) | 0.430 |
| Black | 204 (17.8%) | 88 (16.2%) | 116 (19.4%) |  |
| Other | 83 (7.3%) | 37 (6.8%) | 46 (7.7%) |  |
| Weight (kg) | 82.5 ± 21.9 | 86.1 ±19 | 73.6 ±19.8 | 0.001 |
| BMI (kg/m<sup>2</sup>) | 28.7 ± 7.08 | 27.3 ±5.4 | 28 ±7.3 | 0.191 |
| At Risk for Malnutrition (MST) | 285 (24.9%) | 134 (24.6%) | 151 (25.2%) | 0.695 |
| Clinical Malnutrition | 231 (20.2%) | 118 (21.7%) | 113 (18.9%) | 0.265 |
| Albumin (median [IQR]) | 4 [3.4 - 4.6] | 4 [3.5 - 4.5] | 4 [3.4 - 4.6] | 0.200 |
| Albumin <3.5 g/dL | 172 (15.04%) | 84 (15.4%) | 88 (14.7%) | 0.786 |
| ASA Classification |  |  |  |  |
| 1-2 | 392 (34.3%) | 165 (30.3%) | 227 (37.9%) | 0.007 |
| 3-4 | 751 (65.7%) | 379 (69.7%) | 372 (62.1%) |  |
| Current Smoker | 127 (11.1%) | 65 (11.9%) | 62 (10.4%) | 0.445 |
| Diabetes Mellitus |  |  |  |  |
| Insulin | 89 (7.78%) | 53 (9.7%) | 36 (6%) | 0.008 |
| Non-Insulin | 115 (10.1%) | 64 (11.8%) | 51 (8.5%) |  |
| No | 939 (82.2%) | 427 (78.5%) | 512 (85.5%) |  |
| Hypertension | 529 (46.3%) | 269 (49.4%) | 260 (43.4%) | 0.044 |
| Disseminated Cancer | 144 (12.6%) | 70 (12.9%) | 74 (12.4%) | 0.863 |
| Procedure Type |  |  |  |  |
| Colectomy | 504 (44.1%) | 238 (43.8%) | 266 (44.4%) | 0.225 |
| VHR | 229 (20%) | 105 (19.3%) | 124 (20.7%) |  |
| Pancreatectomy | 204 (17.8%) | 91 (16.7%) | 113 (18.9%) |  |
| Hepatectomy | 155 (13.6%) | 87 (16%) | 68 (11.4%) |  |
| Proctectomy | 51 (4.46%) | 23 (4.2%) | 28 (4.7%) |  |
| Operative Approach |  |  |  |  |
| Laparoscopic | 317 (27.7%) | 141 (25.9%) | 176 (29.4%) | 0.718 |
| Lap converted to Open | 79 (6.9%) | 40 (7.35%) | 39 (6.5%) |  |
| Open | 747 (65.4%) | 363 (66.7%) | 384 (64.1%) |  |
| Surgical Outcomes |  |  |  |  |
| Any Complication | 224 (19.6%) | 113 (20.7%) | 111 (18.5%) | 0.380 |
| Long Length of Stay | 256 (22.4%) | 127 (23.3%) | 129 (21.5%) | 0.435 |

ASA: American Society of Anesthesiologists; MST: Malnutrition Screening Tool; Mean ± standard deviation is reported for continuous features. The frequency and percentage of patients in each category is reported for categorical features.

### Prevalence of Clinical Malnutrition

On admission to the hospital after surgery, 285 (24.9%) patients were found to be “At Risk” for malnutrition using the Malnutrition Screening Tool. A total of 231 patients (20.2%) were diagnosed with clinical malnutrition by a Registered Dietitian. Diagnosis of clinical malnutrition was highest among patients who had pancreatectomy (38.2%) followed by colectomy (23.8%), proctectomy (21.6%), hepatectomy (9.0%) and ventral hernia repair (3.5%).

### Description of Body Composition Differences Between Males and Females

Males and females had significantly different body composition using volumetric CT measures. Histograms depicting the distribution of each imaging feature for males and females separately are depicted in Figure 2. Males had significantly larger muscle size than females for all five muscle groups (Figure 2A: erector spinae: 282.7 ± 66.4 vs. 252.8 ± 55.1 cm^3^/m^2^, lateral abdominus: 342.3 ± 101 vs. 271.8 ± 88 cm^3^/m^2^, psoas: 106.2 ± 28.7 vs. 79.9 ± 21.2 cm^3^/m^2^, quadratus lumborum: 40.4 ± 11.8 vs. 31.3 ± 8.8 cm^3^/m^2^, rectus abdominus: 91.6 ± 30 vs. 72.6 ± 23.1 cm^3^/m^2;^ all p<0.001). Males displayed better muscle quality with higher attenuation for four of the five muscle groups (Figure 2B: erector spinae: 27.3 ± 18.4 vs. 18.5 ± 20 HU, lateral abdominus: 32±17 vs. 26.1± 19 HU, quadratus lumborum: 26±16 vs. 23.4±17 HU, rectus abdominus: 19 ± 17.3 vs. 8 ± 20.9 HU; all p<0.001), which indicates less fatty replacement of muscle tissue compared to females. Males had significantly larger visceral fat depot (1156.6 ± 705 vs. 776.8 ± 493, p<0.001) with lower attenuation (-86.1 ± 9.8 vs. -84.2 ± 9.1, p<0.001), and they had less abdominal subcutaneous fat (1291.8 ± 755 vs. 1868 ± 1020, p<0.001) with greater attenuation (-95.3 ± 9.6 vs. -98.3 ± 9.3 p<0.0001) compared to females (Figure 2B). Principal component analysis (Figure 2C) demonstrated separation between males and females (p=0.001). While the female class was tightly clustered, the distribution of males was more dispersed. The average distance to centroid was greater for males compared to females (3.43 vs. 3.28, p=0.05), reflecting greater variability in the feature measurements.

**Figure 1:**
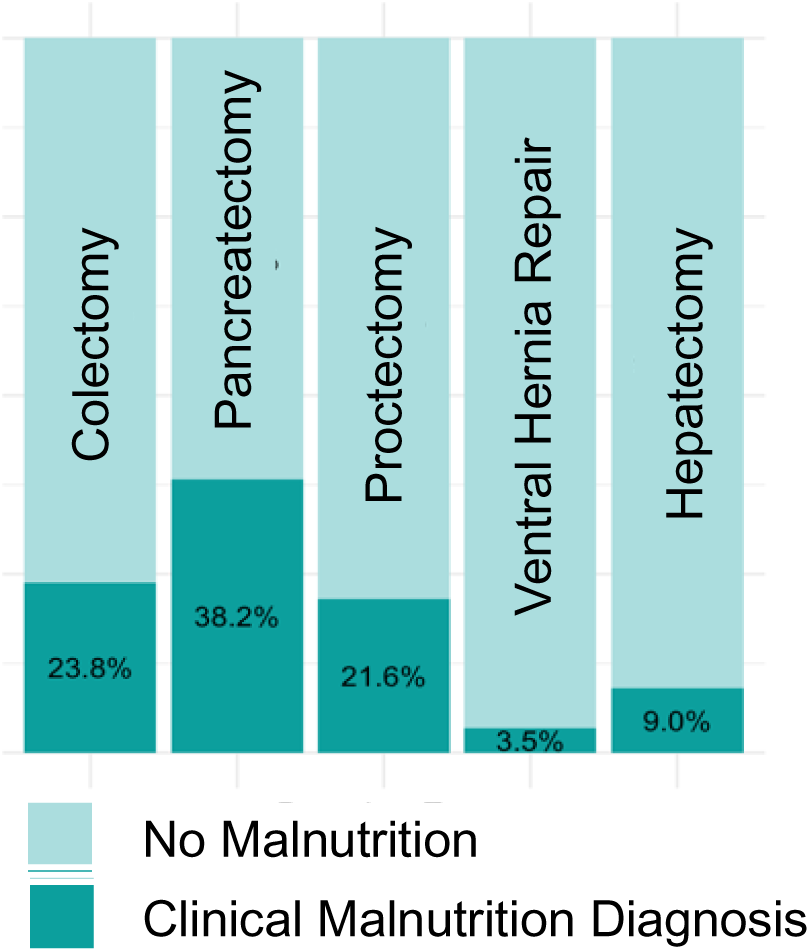
Prevalence of clinical malnutrition varies according to abdominal surgery procedure type.

**Figure 2:**
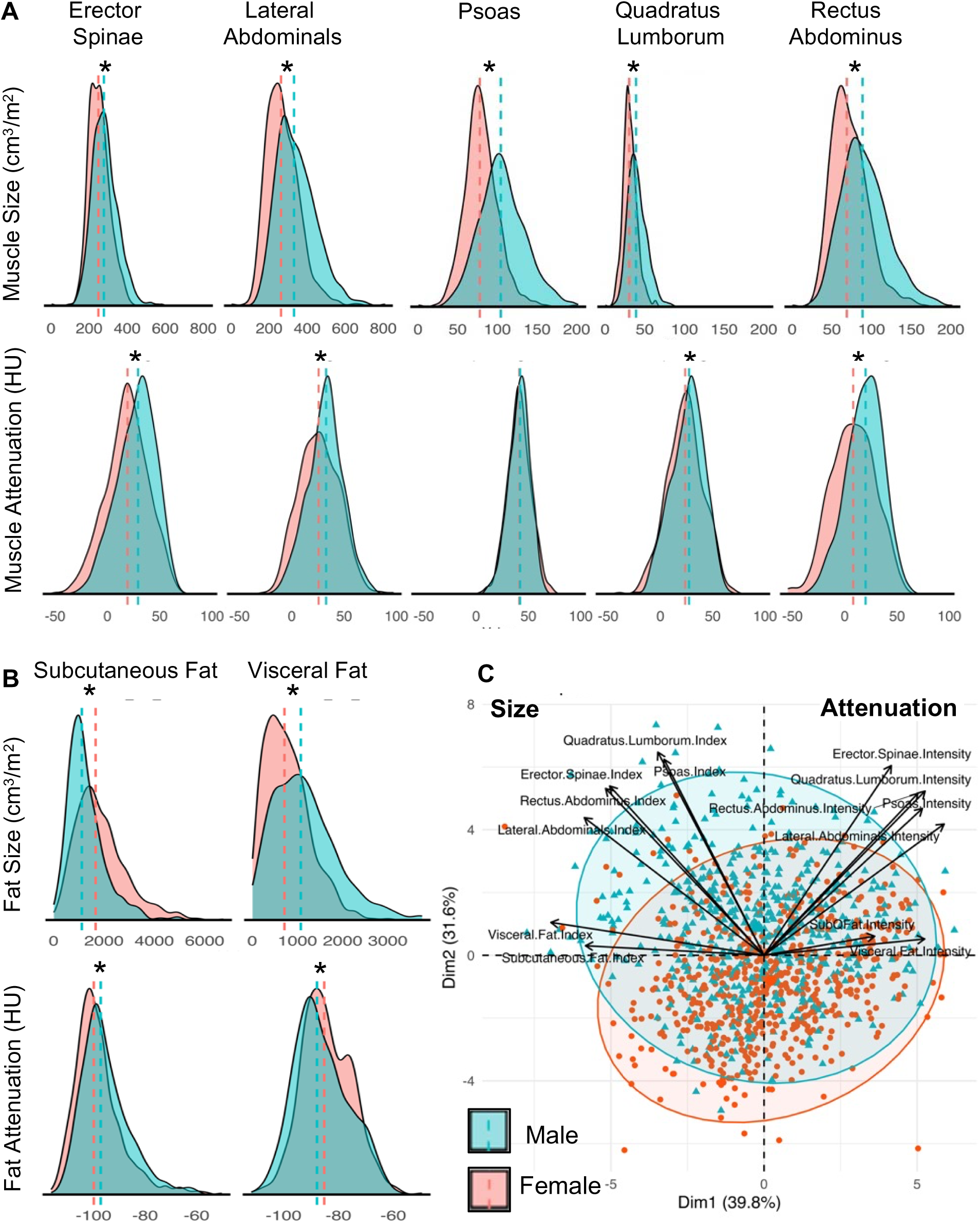
Distribution of imaging features by sex, including size (volume index, cm^3^/m^2^) and attenuation (HU) of (A) individual muscle groups and (B) fat depots. (C) Principal Component Analysis of body composition features cluster according to sex with significantly different centroid and dispersion. *P<0.05

### Average value of Body Composition Features Differ with Malnutrition in Males and Females

Both males and females with clinical malnutrition had significantly decreased size (volume index) for each individual muscle group (psoas, erector spinae, quadratus lumborum, and rectus abdominus, all p<0.001, Figure 3). Females with malnutrition compared to females with normal nutrition had significantly greater fatty replacement (decreased attenuation) for three distinct muscles (erector spinae: 13.1 ± 20 vs. 19.7 ± 20 HU, p<0.001, psoas: 37.7 ± 12 vs 42 ± 11 HU, p<0.0001, quadratus lumborum: 19.9 ± 19 vs. 24.2 ± 17 HU, p<0.01). For males, muscle quality did not differ by nutritional status.

**Figure 3:**
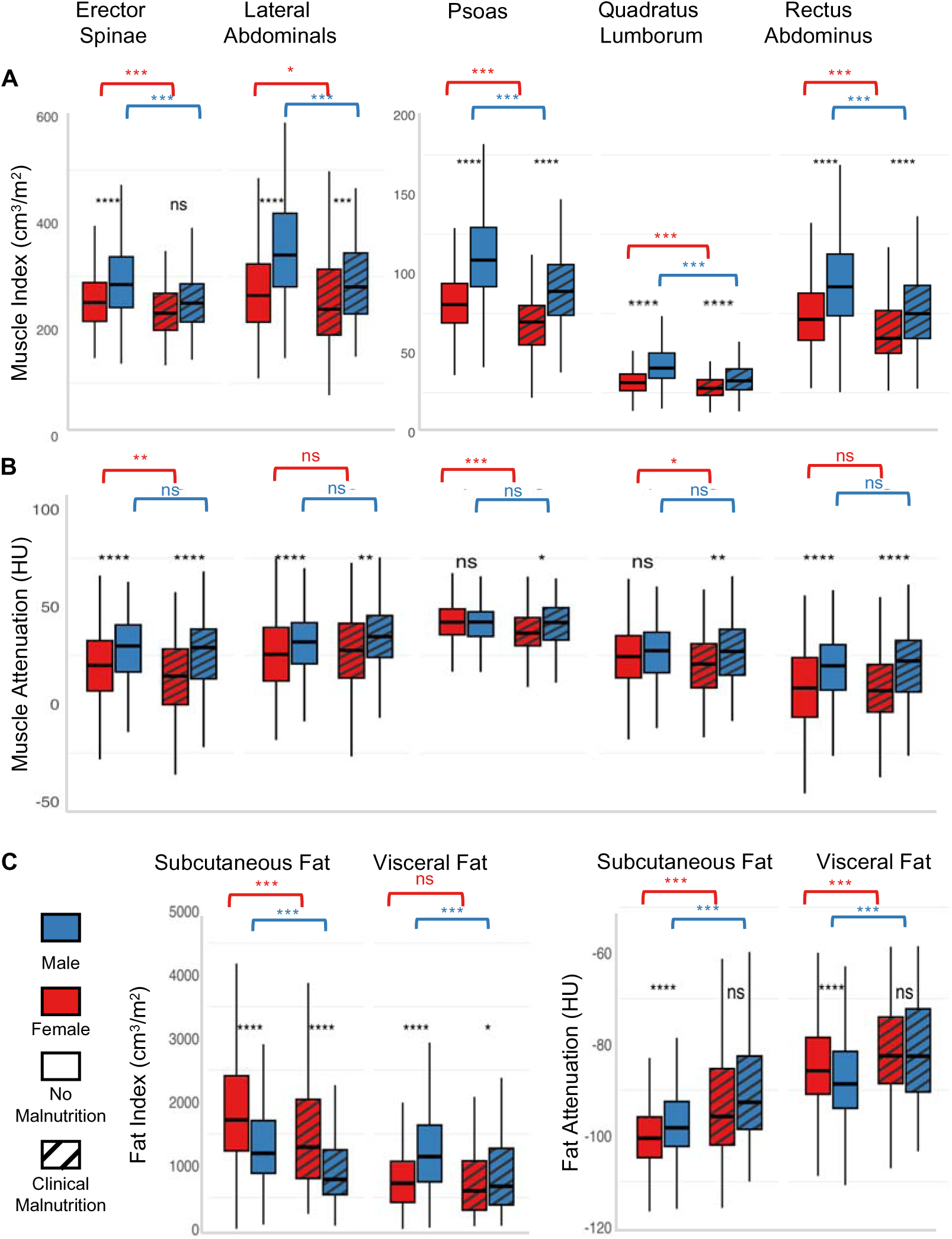
Comparison of each imaging feature (A) muscle volume index, (B) muscle attenuation, and (C) fat volume index & attenuation for males (blue) and females (red) with malnutrition (striped box) and those without (solid). Both males and females with malnutrition have smaller muscle size. Females with malnutrition have significantly decreased attenuation (fatty replacement) for three of five muscle groups. Males do not have significant differences in attenuation by nutrition status. Visceral fat stores decrease significantly in size with malnutrition for males but not for females. Visceral fat attenuation increases for both males and females with malnutrition. Both males and females with malnutrition have significantly smaller subcutaneous fat stores with greater attenuation. *** P<0.0001, ** P<0.001, * P<0.01

Both males and females with malnutrition had significantly smaller subcutaneous fat depot compared to those without malnutrition (Figure 3C). Males with malnutrition had significantly less visceral fat compared to males without malnutrition (869 ± 632 vs. 1236 ± 703 cm^3^/m^2^, p<0.0001). While females were less likely to store visceral fat in general, there was a trend toward loss of visceral fat among females with malnutrition (704 ± 481 vs. 794 ± 495, p=0.076). Both males and females had significantly increased attenuation of subcutaneous and visceral fat, suggesting alteration in size of lipid droplet or fluid content (Figure 3C).

### Association between Body Composition Features and Likelihood of Diagnosis of Clinical Malnutrition

Multivariate logistic regression demonstrated different relationships between individual imaging features and the odds of a diagnosis of clinical malnutrition for males and females (Table 2). For males, smaller psoas (OR 0.59 SDs [0.43 to 0.8], p<0.001), smaller quadratus lumborum (OR 0.59 SDs [0.39 to 0.79], p<0.001), and decreased SMI (OR 0.48 SDs [0.35 to 0.67, p<0.001) were significantly associated with greater likelihood of clinical malnutrition (Table 2a). For females, smaller psoas muscle volume (OR 0.55 SDs [0.41 to 0.74], p<0.0001) and SMI (OR 0.53 SDs [0.39 to 0.73], p<0.001) were significantly associated with increased likelihood of clinical malnutrition.

**Table 2a.**
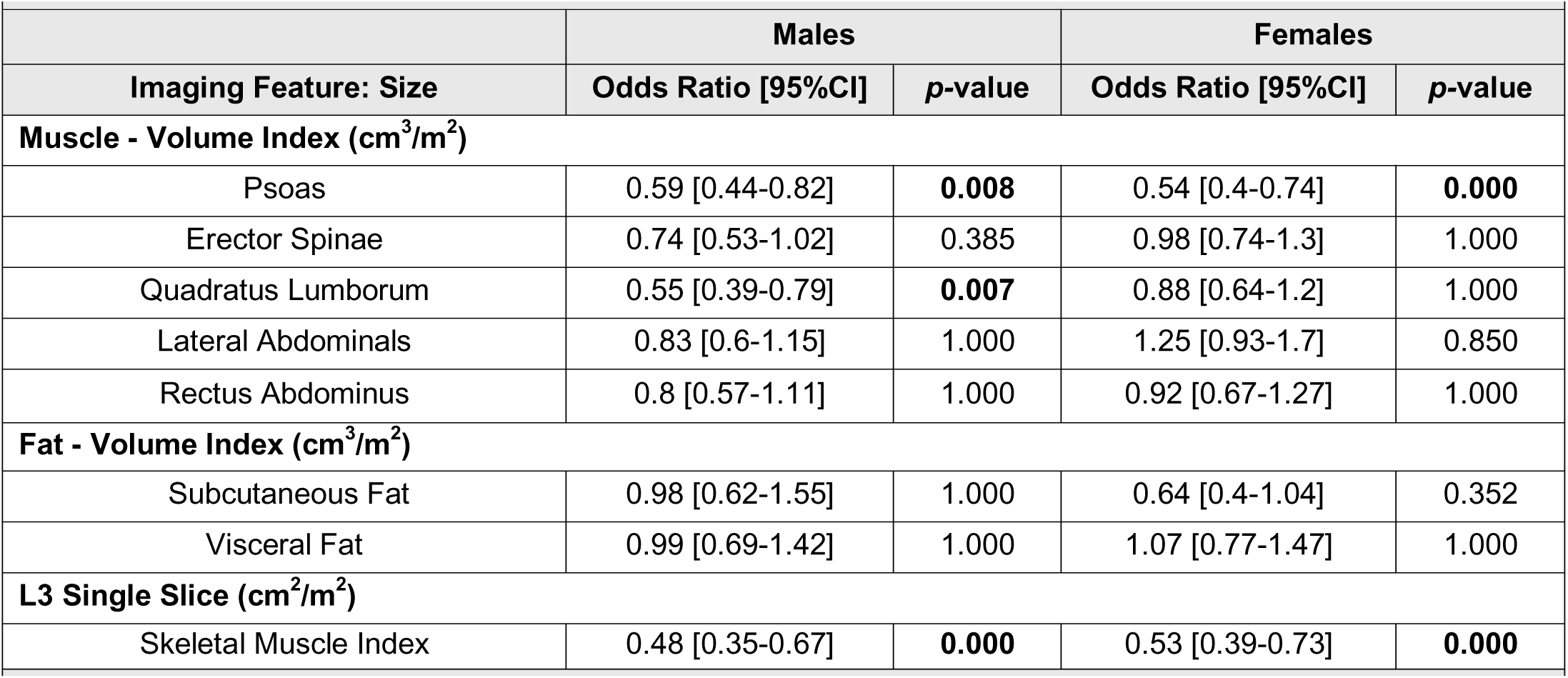
Association between imaging derived feature (size) and likelihood of clinical malnutrition for Males and Females using logistic regression adjusted for age, race, procedure, and smoking.

For males, decreased attenuation (myosteatosis) of erector spinae (OR 0.66 SDs [0.49 to 0.9], p=0.048) and lateral abdominals (OR 0.65 SDs [0.48 to 0.9], p=0.05) was associated with greater risk of malnutrition (Table 2b). For females, decreased attenuation of psoas muscle (OR 0.64 SDs [0.5 to 0.83], p<0.001), erector spinae muscle (OR 0.67 [0.49 to 0.9], p=0.05), and L3 skeletal muscle radiation attenuation (OR 0.67 SD [0.49 to 0.9], p<0.046) was associated with greater odds of malnutrition. For males, increased subcutaneous fat attenuation (decreased size of fat cell) was associated with likelihood of clinical malnutrition (OR 1.51 SDs [1.2 to 1.9], p<0.001). For females, increased subcutaneous fat attenuation (OR 1.73 SDs [1.4 to 2.2], P<0.001) and increased visceral fat attenuation (OR 1.4 SDs [1.11-1.77], p=0.03) were both associated with greater odds of malnutrition.

**Table 2b.**
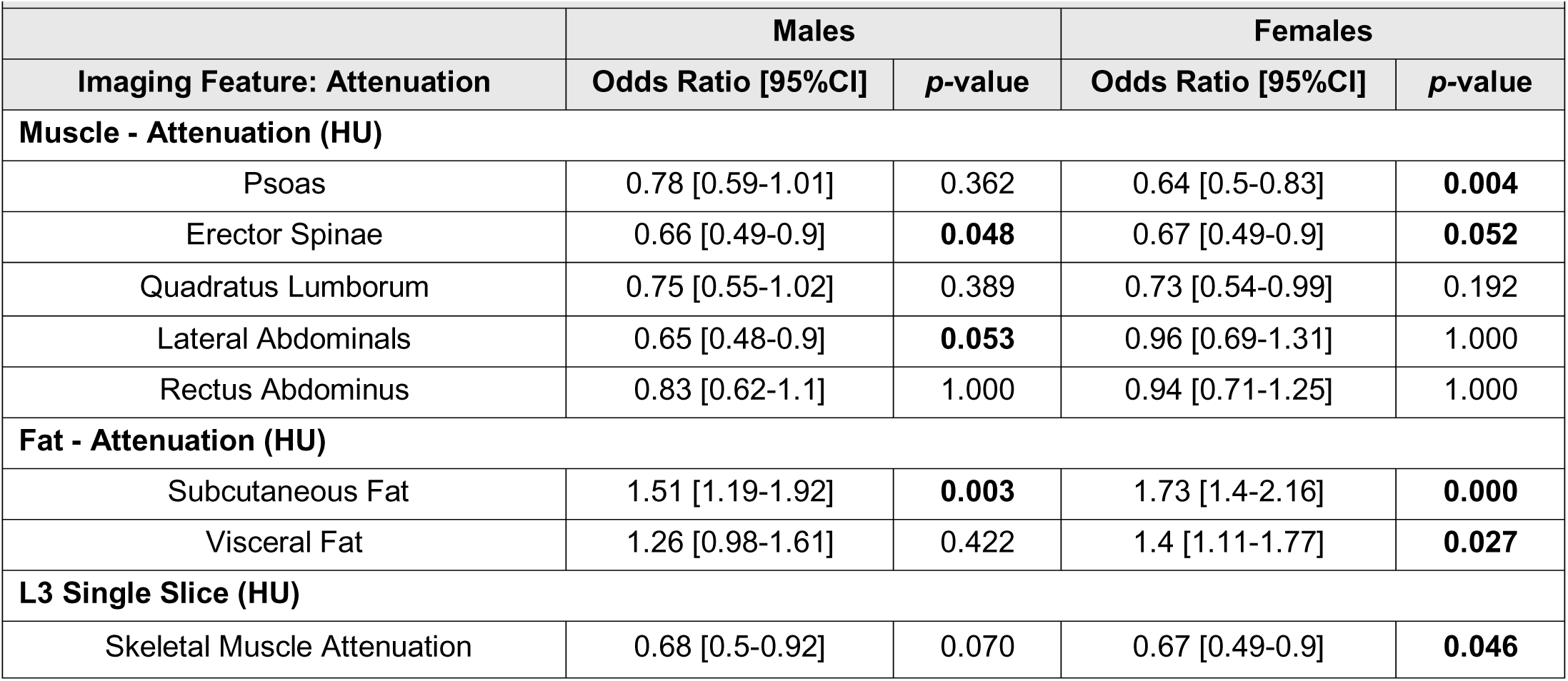
Association between imaging derived feature (attenuation) and likelihood of clinical malnutrition for Males and Females using logistic regression adjusted for age, race, procedure, and smoking.

## DISCUSSION

This study identifies preoperative imaging-derived body composition features associated with clinical malnutrition among males and females. Our findings demonstrate that body composition analysis using routine preoperative CT scans can identify various characteristics (Imaging-Derived Phenotypes) predictive of malnutrition. Further, while both males and females with malnutrition show reduced muscle size, the patterns of muscle quality deterioration (myosteatosis) and fat distribution vary significantly between sexes, suggesting that accounting for patient sex and multiple muscle and fat metrics may be necessary for accurate nutritional assessment using imaging-based body composition analysis.

Several key findings emerge from our analysis. First, in keeping with the literature, we observed that males and females exhibit distinct body composition profiles independent of nutritional status, with males showing greater volumes of both muscle and visceral fat, while females have smaller muscle with more subcutaneous fat deposition. Both males and females with malnutrition exhibited significantly smaller muscle volumes compared to those without malnutrition, consistent with known associations between sarcopenia and malnutrition. (40) Importantly, females with normal nutritional status had similar muscle size compared to males with malnutrition, which highlights the discrepancy in size between the sexes and the importance of sex-disaggregated analysis. Consistent with previous findings, the relationship between malnutrition risk and decreased muscle size—both psoas volume index and L3 SMI—was true for both sexes. However, the association between other muscle features and malnutrition for males and females separately suggest that a more nuanced approach using comprehensive volumetric analysis is needed to develop imaging biomarkers and for future use in predictive risk models.

Second, we found that females with malnutrition had significantly decreased mean muscle attenuation for three out of the five muscle groups compared to females without malnutrition. For males, however, there was no significant difference in mean muscle attenuation for males with or without malnutrition. Decreased muscle attenuation is seen with myosteatosis and represents decreased muscle quality. Myosteatosis includes both intramuscular adipose deposition and intramyocellular lipid droplet accumulation. Intramuscular adipose tissue has been linked to impaired skeletal muscle function with decreased strength and metabolic disease, including insulin resistance. (41, 42) Using logistic regression, we determined that differences in muscle attenuation were valuable for determining likelihood of malnutrition in both males and females. In a recently published study, Xie et al. also found sex-specific differences in myosteatosis among older hospitalized females with malnutrition. This relationship was not significant for males. Together, these similar findings suggest that fatty deposition and change in muscle quality are more sensitive imaging characteristics for females with malnutrition. (43) Increased fatty replacement, specifically intramuscular adipose tissue, has been associated with decreased force generation 1and worse function, especially among those with metabolic disease.(41) Similarly, myosteatosis is associated with worse outcomes after surgery. (44-46) For females, who inherently have smaller muscle size, increased fatty replacement may be a more significant marker of decreased muscle function as seen in sarcopenia and frailty. These imaging findings are clinically meaningful; in a meta-analysis of 19 papers, preoperative myosteatosis has been shown to be associated with poor outcomes and decreased overall survival after surgery for gastrointestinal malignancy. (47) These findings highlight the value of incorporating muscle quality in addition to muscle size as an indicator of malnutrition, especially for female patients.

Our findings regarding fat depletion and change in attenuation are particularly noteworthy. Males are known to have greater visceral fat deposition due to hormonal influences. (22, 23, 48) Here we demonstrate an association between clinical malnutrition and loss of visceral fat with increased attenuation, which has potential to be an imaging-based biomarker for inadequate nutrition. CT findings demonstrate both loss of stored fat and decreased size of lipid droplet possibly secondary to increased lipolysis and enhanced fatty acid uptake which occurs as a compensatory response to inadequate nutrition. (13, 49-51). Adipocyte lipolysis is regulated by multiple hormonal cues that including insulin and catecholamines known to be aberrant in malnutrition. The amount of visceral fat was not significantly different for females with and without malnutrition. However, increased mean attenuation of both subcutaneous and visceral fat, which may represent lipolysis and fat mobilization, was associated with greater likelihood of malnutrition. In non-human primates and in studies of cancer cachexia, increased adipose attenuation has been linked to increased mortality. (52) Change in adipose tissue attenuation is thought to reflect adipose remodeling, including decreased fat cell volume with increased fibrosis of surrounding connective tissue. (53, 54) This imaging characteristic of increased adipose attenuation has been associated with deteriorating health and is an imaging marker of former obesity, both in weight loss associated with aging and as seen in cancer-cachexia. (52, 54-57) Both males and females with malnutrition had increased attenuation of visceral and subcutaneous fat suggesting that after depletion of stored lipids, lipocytes reach a similar minimum size in both males and females. These imaging-based findings have important implications for nutritional assessment protocols, can serve as imaging biomarkers for underlying physiologic response to inadequate nutrition, and will improve our ability to identify malnutrition in a high-throughput and reliable way.

These results have several important clinical implications. First, they suggest that nutritional assessment tools incorporating body composition analysis should account for sex-specific differences in both muscle and fat characteristics. Second, the finding that muscle quality is particularly important in females indicates that evaluation of muscle mass alone may be insufficient for accurate nutritional assessment. Third, the comprehensive volumetric analysis approach used in this study provides a more thorough assessment than traditional single-slice measurements and supports the development of image-based screening models to identify patients with malnutrition in the preoperative setting, presenting an opportunity for preoperative nutritional optimization.

Several limitations should be considered when interpreting these results. First, the retrospective nature of the study and single-institution setting may limit generalizability. Second, while our cohort included a diverse range of abdominal surgeries, procedure-specific analyses were limited by sample size in some categories. Third, the use of clinical malnutrition diagnosis as the reference standard, while pragmatic, may be subject to subjectivity despite standardized criteria. Finally, the cross-sectional nature of our analysis prevents determination of causality in the relationship between body composition changes and malnutrition.

Future research should focus on validating these findings in prospective, multi-center studies and investigating whether sex-specific body composition analysis can improve the accuracy of preoperative nutritional risk assessment. Additionally, studies examining the longitudinal changes in body composition features during nutritional intervention may provide insights into sex-specific responses to nutritional support.

## CONCLUSION

This study demonstrates significant associations between body composition features on preoperative imaging and malnutrition among males and females undergoing abdominal surgery. These findings suggest that sex-specific approaches to body composition analysis may improve the accuracy of nutritional assessment and highlight the potential value of comprehensive volumetric analysis over traditional single-slice measurements. Implementation of these findings could lead to more precise and personalized approaches to preoperative nutritional assessment and intervention strategies.

## Data Availability

All data produced in the present study are available upon reasonable request to the authors

## Abbreviations

BMI: Body Mass Index
CT: Computed Tomography
HU: Hounsfield Units
SMI: Skeletal Muscle Index
SMRA: Skeletal Muscle Radiation Attenuation
L3: 3^rd^ Lumbar Vertebrae
AAIM: Academy ASPEN Indicators of Malnutrition
NSQIP: National Surgery Quality Improvement Program
OR: Odds Ratio

## Author Contributions

Conception and study design: Gershuni, Luks, Compher, Witschey, Wu

Data acquisition: Gershuni, Luks, Damani, Witschey, Wu

Data analysis and interpretation: Damani, Vasisht, Compher, Tasian, Titchenell, Li, Wu, Gershuni

Writing – drafting the article: Gershuni, Damani, Vasisht, Witschey, Wu

Writing – critical review & editing: Gershuni, Vargas, Titchenell, Compher, Tasian, Wu, Witschey

## Acknowledgements

We wish to acknowledge with gratitude the Penn Clinical Nutrition Support Service Registered Dietitians for their instrumental role in collecting the data that underpins this research. Their expertise and dedication were vital in gathering comprehensive and accurate information necessary for our study. We would like to express our sincere gratitude to James Rowe, RN (Surgical Clinical Reviewer for NSQIP) for his valuable contribution in data collection and abstraction. His meticulous work was essential to the successful completion of this study. Special thanks to Jeffrey Duda for imaging data acquisition and Yue Ren for assistance with initial data cleaning and processing, which established a solid foundation for our analysis. Our sincere thanks go to Valerie Luks, MD for data collection and initial data exploration. Furthermore, we are grateful to Rachel Kelz, MD MBA MSCE FACS for her guidance and support throughout this research project. Her expertise and mentorship significantly enhanced the quality of this work.

---

**Supplementary Table 1.** List of imaging-derived body composition features.

|  | Size Feature | Attenuation Feature |
| --- | --- | --- |
| 2D | L3 Skeletal Muscle Index<br>(cm<sup>2</sup>/m<sup>2</sup>) | L3 Skeletal Muscle<br>Radiation Attenuation (HU) |
| 3D | Psoas Index (cm<sup>3</sup>/m<sup>2</sup>) | Psoas Intensity (HU) |
| 3D | Quadratus Lumborum<br>Index (cm<sup>3</sup>/m<sup>2</sup>) | Quadratus Lumborum<br>Intensity (HU) |
| 3D | Erector Spinae Index<br>(cm<sup>3</sup>/m<sup>2</sup>) | Erector Spinae Intensity<br>(HU) |
| 3D | Rectus Abdominus Index<br>(cm<sup>3</sup>/m<sup>2</sup>) | Rectus Abdominus<br>Intensity (HU) |
| 3D | Lateral Abdominals Index<br>(cm<sup>3</sup>/m<sup>2</sup>) | Lateral Abdominals<br>Intensity (HU) |
| 3D | Visceral Fat Index (cm<sup>3</sup>/m<sup>2</sup>) | Visceral Fat Intensity (HU) |
| 3D | Subcutaneous Fat Index<br>(cm<sup>3</sup>/m<sup>2</sup>) | Subcutaneous Fat Intensity<br>(HU) |

## Précis

Males and females exhibit distinct body composition patterns in malnutrition. While both sexes show reduced muscle mass with malnutrition, only females demonstrate significant myosteatosis. Sex-specific associations between CT-derived body composition features and clinical malnutrition highlight the need for sex-specific approaches in nutritional assessment of surgical patients.

## Notes

**Funding:** Research reported in this publication was supported by the National Center for Advancing Translational Sciences of the National Institutes of Health under award numbers KL2TR001879 and UL1TR001878. The content is solely the responsibility of the authors and does not necessarily represent the official views of the National Institutes of Health. This work was also supported in part by the Institute for Translational Medicine and Therapeutics’ (ITMAT) Transdisciplinary Program in Translational Medicine and Therapeutics, the McCabe Fund Award, and the Biomedical Data Sciences Core of the Center for Molecular Studies in Digestive and Liver Diseases (P30 DK050306), and the Penn Center for Nutritional Science and Medicine. Additional support was provided by NIH grants R01 DK139663, R01 HL171709, P41 EB029460, and R01 HL169378. The funding sources had no role in the design, execution, interpretation, or writing of the study.

### Competing Interest Statement

The authors have declared no competing interest.

### Funding Statement

Research reported in this publication was supported by the National Center for Advancing Translational Sciences of the National Institutes of Health under award numbers KL2TR001879 and UL1TR001878. The content is solely the responsibility of the authors and does not necessarily represent the official views of the National Institutes of Health. This work was also supported in part by the Institute for Translational Medicine and Therapeutics' (ITMAT) Transdisciplinary Program in Translational Medicine and Therapeutics, the McCabe Fund Award, and the Biomedical Data Sciences Core of the Center for Molecular Studies in Digestive and Liver Diseases (P30 DK050306), and the Penn Center for Nutritional Science and Medicine. Additional support was provided by NIH grants R01 DK139663, R01 HL171709, P41 EB029460, and R01 HL169378. The funding sources had no role in the design, execution, interpretation, or writing of the study. 

### Author Declarations

Ethics committee/IRB of Perelman School of Medicine at University of Pennsylvania waived ethical approval for this work

